# Region-specific cerebral blood flow differentiates cognitively impaired and unimpaired individuals with core Alzheimer’s disease pathology

**DOI:** 10.64898/2026.08.22.26361093

**Authors:** Sara Fernandes-Taylor, Ira Driscoll, Matthew Glittenberg, Brianne Breidenbach, Adam Paulsen, Katelyn Hauge, Avery Rhodes, Leonardo A. Rivera-Rivera, Laura Eisenmenger, Kevin M. Johnson, Aaron Field, Catherine Gallagher, Sterling C. Johnson, Sanjay Asthana, Mark Sager, Cynthia Carlsson, Barbara Bendlin, Bradley Christian, Tobey Betthauser, Caitlin S. Latimer, Ozioma C. Okonkwo

## Abstract

**Background:** Up to 30% of individuals harboring core Alzheimer’s disease (AD) neuropathology—amyloid-β (Aβ;A) and tau (T)—remain cognitively unimpaired.

**Objective:** We examine whether MRI measures of cerebral blood flow (CBF) and white matter hyperintensities (WMH) differentiate cognitively impaired (AD or mild cognitive impairment (MCI);AD+MCI) from non-demented individuals with (NDAN) and without (Controls) core AD neuropathology.

**Methods:** A retrospective cohort study (2018-2023) using data from The Wisconsin Registry for Alzheimer’s Prevention and Wisconsin Alzheimer’s Disease Research Center studies employed linear models comparing group differences in regional gray matter (GM) CBF and whole brain WMH volume. Participants (N=500) underwent 3T MRI, Aβ-(^11^C-Pittsburgh Compound B) and tau-(^18^F-MK6240) PET, and neuropsychological assessments. We categorized participants based on PET and cognitive status as Controls (n=416; cognitively unimpaired, A-T-), NDAN (n=38; cognitively unimpaired, A+T+), or AD+MCI (n=46; cognitively impaired, A+T+).

**Results:** Participants were 67 years old (mean), 68% female, and 37% *APOE* ε4+. NDAN, like Controls, had significantly higher CBF than AD+MCI in GM regions susceptible to neurofibrillary tangle formation in early AD (frontal and temporal cortices, and limbic regions (*p*s<0.05)). Largest CBF differences (22-34%) were observed in GM regions that accumulate Aβ early but remain largely tangle-free during early AD progression, namely the occipital and parietal cortices. WMH volume differentiated AD+MCI from Controls (*p*=0.002) but not NDAN (*p*=0.16).

**Conclusion:** CBF differentiated NDAN from AD+MCI, highlighting vascular contributions to cognitive resilience. Most pronounced CBF preservation in NDAN was observed in parietal and occipital cortices, regions typically free of tau until late stages of AD progression.

## BACKGROUND

Between 12-30% of older individuals accumulate significant core Alzheimer’s disease (AD) pathology without presenting with clinically relevant cognitive deficits.^1–4^ These individuals who are non-demented (with) AD neuropathology (NDAN) represent a phenotype of cognitive resilience. Three recent reviews describe neuronal, neurodegenerative, and inflammatory processes that differentiate NDAN^1,5,6^ but discuss few vascular characteristics. Although vascular contributions, including blood-brain barrier maintenance^7,8^ and white matter microstructural integrity,^9,10^ to resilience against AD-associated cognitive decline have been identified, it remains unknown whether they differ in NDAN.

The vascular hypothesis of AD^11^ suggests that altered cerebral blood flow (CBF) is an early and intrinsic part of AD progression.^12–14^ This phenomenon first manifests as a brief period of hyperperfusion in the preclinical stages of the disease, potentially as a compensatory mechanism aimed at preserving metabolic support and oxygen delivery as AD pathology, namely amyloid-β (Aβ) plaques and neurofibrillary tangles (tau), accumulates.^15–19^ Subsequently, a reduction in CBF is observed with disease progression,^12,19^ specifically in regions associated with the deposition of tau, including parietal cortices, the hippocampus, and the temporal lobes in general,^20,21^ and the hypoperfusion intensifies with AD progression.^12,15,19^ Lower CBF is thought to result from a combination of vascular changes, particularly small vessel disease, including endothelial dysfunction and amyloid angiopathy, which disrupt normal blood flow to neurons and contribute to neuronal dysfunction and neurodegeneration.^22^ The extent to which CBF alterations are evident in NDAN remains unknown.

White matter hyperintensities (WMH) are also thought to result from small vessel pathology and contribute to cognitive decline.^10^ WMH, areas of white matter damage resulting from chronic cerebrovascular disease, are associated with worse cognitive outcomes and may exacerbate the clinical expression of AD when core neuropathology is present.^9,10^ Conversely, the preservation of white matter microstructure is associated with resilience to AD, specifically cognitive resilience to tau accumulation.^23,24^ To date, it remains unclear whether WMH contribution to cognitive decline differentiates NDAN from those with AD or mild cognitive impairment (MCI).

The aim of the present study is to investigate whether cerebral hemodynamics and WMH contribute to cognitive resilience in AD. Here we compare CBF and WMH in cognitively unimpaired individuals free of (Controls) or harboring (NDAN) core AD neuropathology relative to AD+MCI.

## METHODS

### Participants

Data of interest was available for five hundred participants from the Wisconsin Registry for Alzheimer’s Prevention (WRAP)^25^ and the Wisconsin Alzheimer’s Disease Research Center (WADRC).^26^ The WRAP and WADRC are two prospective cohorts enriched for parental history of AD at enrollment. Participants underwent neuropsychological assessments, magnetic resonance imaging (MRI), and positron emission tomography (PET) for Aβ and tau. All procedures were approved by the University of Wisconsin Health Sciences Institutional Review Board, and all individuals signed informed consent prior to participation. Researchers may obtain access to de-identified data used for this study by request and committee approval.

### Cognitive Status Classification

Consensus diagnoses of MCI or probable AD were assigned based on the National Institute on Aging-Alzheimer’s Association (NIA-AA) criteria.^25,27–29^ Participants who did not meet criteria for MCI or AD (AD+MCI; CBF: n=32;WMH: n=46), were deemed cognitively unimpaired [CU; CBF: n=330, WMH: n=454).

### PET Imaging

PET imaging was performed on a Siemens EXACT HR+ or a Siemens Biograph Horizon scanner. Imaging methods including radio-pharmaceutical production, image reconstruction, acquisition protocols, processing and quantification of PET are detailed elsewhere.^30–32^ Parametric images were co-registered to T1-weighted MRI.

To quantify Aβ load, distribution volume ratio (DVR) maps were generated from a 70-min dynamic PiB PET acquisition using Logan graphical analysis with cerebellar gray matter as reference. Participants were Aβ positive (A+) or negative (A-) based on a previously published global cortical distribution volume ratio (DVR) threshold of >1.19.^32^

For tau, standardized uptake value ratios (SUVRs) were calculated from a 20-min dynamic MK6240 PET acquisition, 70 min post bolus injection using the inferior cerebellar grey matter as reference.^30^ Participants were characterized as tau positive (T+) or negative (T-) based on visual reads of volume-weighted mean SUVRs calculated for Harvard-Oxford atlas regions associated with neuropathological Braak staging (Braak III: temporal portion of fusiform gyrus; Braak IV: inferior and middle temporal gyri and insular cortex; Braak V: superior temporal, angular, supramarginal, and middle frontal gyri, planum temporale and the occipital portion of the fusiform gyrus; Braak VI: Heschl’s gyrus, intracalcarine cortex, amygdala, posterior cingulate, precuneus, lingual gyrus, caudate, and putamen).^33^ Participants were assigned a Braak score based on the highest Braak stage composed of regions showing MK6240 uptake *in vivo*, reflecting the spatial spread of tau pathology described in the neuropathological literature.

Specifically, a consensus panel of a neuroradiologist and neuroimaging experts blinded to clinical and amyloid status of the participant determined Braak stage using a structured classification based on visual ratings of regional patterns of MK6240 uptake by comparing reference distributions derived from young, cognitively unimpaired adults serving as controls. Detailed methods for determining *in vivo* Braak staging based on tau-PET are published elsewhere.^30,34^

### Participant Classification

For analysis purposes, CU participants were classified as either Controls (CBF: n=299; WMH: n=416) if they were A-T- or NDAN (CBF: n=31; WMH: n=38) if A+T+ (Braak III+). AD+MCI (CBF: n=32; WMH: n=46) had a consensus diagnosis of cognitive impairment (probable AD or MCI) and were A+T+ (Braak III+).

### Magnetic Resonance Imaging

Participants underwent 3D MRI on either 3T GE x750 or GE Signa Premier scanner using SCAN-compliant T1 and T2-FLAIR protocols.^35^ Scanning was completed after a 4-hour fast from food, caffeine, tobacco, and vasomodulatory medications. T1-weighted images were bias-corrected, tissue class segmented, and spatially normalized to MNI152 standard space.^36,37^

### CBF Protocol

Resting CBF was acquired with background-suppressed pseudo-continuous arterial spin labeling (pCASL)^38,39^ with a 3-D fast spin echo spiral sequence using a stack of variable-density spiral 4 ms-readout and 8 interleaves, which demonstrated excellent test-retest reliability (r > 0.95).^40,41^ Scan parameters included TE/TR = 10.5 ms/4.9 s, FOV = 240mm, matrix size = 128 × 128, NEX = 3, and labeling RF amplitude = 0.24mG. For CBF flow quantitation and image registration, the sequence included a proton density (PD) acquisition with the same imaging sequence/image slab location. The pCASL sequence was acquired in under 5 min.

CBF images were processed using SPM12 by registering each subject’s co-localized PD image to their T1-weighted image, applying the derived transformation matrix to the average quantitative CBF map, then spatially normalizing the T1-weighted and associated CBF image to the MNI template, with resampling to a 2 × 2 × 2 mm voxel size. The normalized CBF maps were then smoothed using an 8-mm FWHM (Full-Width at Half-Maximum) Gaussian kernel. Inter-individual variations in global perfusion were corrected by scaling each voxel in the map by the mean whole brain CBF. CAT12 was used to segment the T1 images and to estimate the registration parameters to transform the probability maps to MNI152NLin2009cAsym space. The resulting maximum probability map was modified by filling remaining holes with median value and applying brainmask_T1.nii to remove non-brain areas. Labels in native space were derived from the “MICCAI 2012 Grand Challenge and Workshop on Multi-Atlas Labeling” (http://www.neuromorphometrics.com/2012_MICCAI_Challenge_Data.html), which were created using data from 35 subjects released under the Creative Commons Attribution-NonCommercial from the OASIS project (http://www.oasis-brains.org/) and Neuromorphometrics, Inc. (http://Neuromorphometrics.com/) under academic subscription. We examined CBF in gray matter regions associated with neurofibrillary tangle formation throughout the AD disease course given the demonstrated associations between tau and both cognitive decline and vascular injury.^42–44^

### WMH Protocol

Total WMH volume was calculated using the Lesion Growth Algorithm (LGA) in the Lesion Segmentation Tool version 1.2.2 in SPM12. This high reliability, automated segmentation toolbox is open source and uses T1-weighted and T2-FLAIR images for lesion segmentation.

Lesions are seeded based on spatial and intensity probabilities from T1 images and hyperintense outliers on T2-FLAIR images. The initial threshold (0.30) created the binary conservative lesion belief map from the gray and white matter lesion belief maps. A growth algorithm then grew the seeds from the conservative lesion belief map toward a probabilistic liberal lesion belief map from gray matter, white matter, and cerebrospinal fluid (CSF) belief maps. A threshold value of 1.00 on the resulting lesion belief map removed voxels with a low probability of being a lesion.

Based on published consensus standards^45^, all final segmentation maps were visually inspected by a team blind to participants’ other characteristics, including amyloid and diagnostic status. When there was a quality control concern, the results were adjudicated by a radiologist.

### Statistical Analyses

Statistical analyses were conducted in Stata v. 18.0.^46^ Sample characteristics were compared using non-parametric Mann-Whitney U and Wilcoxon rank sum tests between the three groups because group sizes were relatively small. Linear models compared outcome variables (CBF and WMH lesion volume) between the groups. Models were adjusted for age, sex and *APOE* ε4 allele carriage (*APOE*4+). CBF models were additionally adjusted for total gray matter volume and days between CBF measurement and tau PET measurement. WMH models were additonally adjusted for total intracranial volume, calculated as the sum of total white matter, total gray matter and CSF volumes. Dependent variables were skewed and thus log-transformed prior to multivariable comparisons. Huber-White robust standard errors were calculated. *P*-values were adjusted for multiple comparisons using the Benjamini-Hochberg method to account for non-independence among outcomes.

## RESULTS

### Sample Characteristics

Charateristics of the entire sample with available CBF data and and group differences are summarized in Table 1. The CBF sample was on average 67 years old and college-educated, 91% were white, 68% female, 37% *APOE*4+, and 71% had a parental history of AD. Controls were significantly younger than either NDAN or AD+MCI (all *p*s < 0.01), and NDAN were significantly younger than AD+MCI (*p* = 0.02). Both AD+MCI and NDAN had a significantly higher percentage of *APOE*4+ compared to Controls (*p* < 0.001). Parental history of AD was more prevalent in NDAN than both the Controls or AD+MCI (all *p*s < 0.01). Time intervals between CBF and tau-PET scan did not significantly different between groups.

**Table 1.** Background characteristics of the sample with available cerebral blood flow (CBF) measures.

|  | Group |  |  |  | Between-group Comparisons<br>(*p*-value) |  |  |
| --- | --- | --- | --- | --- | --- | --- | --- |
|  | Entire Sample | AD+MCI | NDAN | Controls | NDAN vs<br>AD+MCI | NDAN vs<br>Controls | AD+MCI<br>vs Controls |
| **No. (%)** | 362 (100.0%) | 32 (8.8%) | 31 (8.6%) | 299 (82.6%) |  |  |  |
| **Age, mean (SD)** | 67.0 (7.6) | 74.3 (6.0) | 70.2 (6.5) | 65.9 (7.64) | **0.02** | **0.002** | **<0.001** |
| **APOE4+, No. (%)** | 134 (37%) | 23 (72%) | 20 (63%) | 90 (30%) | 0.53 | **<0.001** | **<0.001** |
| **Education (years), mean (SD)** | 16.1 (2.3) | 16.0 (2.4) | 16.0 (2.6) | 16.2 (2.2) | 0.72 | 0.94 | 0.65 |
| **Female, No. (%)** | 246 (68%) | 17 (53%) | 23 (74%) | 206 (68%) | 0.14 | 0.54 | 0.07 |
| **Parental History of AD, No. (%)** | 259 (71%) | 20 (63%) | 30 (97%) | 209 (70%) | **0.001** | **0.002** | 0.39 |
| **Race, No. (%)** |  |  |  |  |  |  |  |
| American Indian | 10 (3%) | 1 (3%) | 1 (3%) | 8 (3%) | 1.00 | 0.86 | 0.88 |
| Asian | 0 (0%) | 0 (0%) | 0 (0%) | 0 (0%) | NA | NA | NA |
| Black or African American | 20 (6%) | 1 (3%) | 2 (7%) | 17 (6%) | 0.98 | 0.86 | 0.54 |
| Other | 3 (1%) | 0 (0%) | 0 (0%) | 3 (1%) | 1.00 | 0.58 | 0.57 |
| White | 329 (91%) | 30 (94%) | 28 (90%) | 271 (91%) | 0.97 | 0.95 | 0.56 |
| **Difference between CBF & tau-PET scans (days), mean (SD)** | 534.3 (566.5) | 540.5 (579.2) | 498.5 (628.5) | 537.3 (560.3) | 0.30 | 0.23 | 0.87 |

**Bolded *p*-values** denote results significant at $p < 0.05$ .
**Note:** Between group comparisons are Mann-Whitney U and Wilcoxon rank sum tests.
**Abbreviations:** AD+MCI: Alzheimer's Disease or Mild Cognitive Impairment, APOE4+: carrier of one or both Apolipoprotein E ε4 alleles, CBF: Cerebral Blood Flow, NDAN: Non-demented Alzheimer's neuropathology, SD: Standard Deviation, tau-PET: MK6240 positron emission tomography.

Group charateristics and differences in the WMH sample are summarized in Table 2. The WMH sample was on average 67.2 years of age and college-educated, 90% white, 68% female, 37% *APOE*4+ and 73% had a parental history of AD. Controls were significantly younger than both NDAN and AD+MCI (all *p*s < 0.001). Both AD+MCI and NDAN had a significantly higher percentage of *APOE*4+ compared to Controls (all *p*s < 0.001). Parental history of AD was more prevalent in NDAN than in Controls or AD+MCI (all *p*s < 0.05).

**Table 2.** Background characteristics of the sample with available white matter hyperintensity (WMH) measures.

|  | Group |  |  |  | Between-group Comparisons<br>(*p*-value) |  |  |
| --- | --- | --- | --- | --- | --- | --- | --- |
|  | Entire<br>Sample | AD+MCI | NDAN | Controls | NDAN vs<br>AD+MCI | NDAN vs<br>Controls | AD+MCI<br>vs Controls |
| **No. (%)** | 500 (100%) | 46 (9%) | 38 (8%) | 416 (83%) |  |  |  |
| **Age**, mean (SD) | 67.2 (7.3) | 73.3 (5.7) | 70.7 (6.0) | 66.2 (7.1) | 0.08 | **<0.001** | **<0.001** |
| **APOE4+**, No. (%) | 185 (37%) | 31 (67%) | 25 (66%) | 129 (31%) | 0.88 | **<0.001** | **<0.001** |
| **Education (years)**, mean (SD) | 16.1 (2.3) | 16.8 (2.4) | 16.3 (2.6) | 16.1 (2.3) | 0.16 | 0.26 | 0.53 |
| **Female**, No. (%) | 340 (68%) | 28 (61%) | 30 (79%) | 283 (68%) | 0.08 | 0.16 | 0.33 |
| **Parental History of AD**, No. (%) | 366 (73%) | 34 (74%) | 37 (97%) | 295 (71%) | **0.003** | **<0.001** | 0.67 |
| **Race**, No. (%) |  |  |  |  |  |  |  |
| American Indian | 12 (2%) | 1 (2%) | 1 (3%) | 10 (2%) | 0.89 | 0.93 | 0.92 |
| Asian | 1 (0%) | 0 (0%) | 0 (0%) | 1 (0%) | NA | 0.76 | 0.74 |
| Black/African American | 34 (7%) | 2 (4%) | 3 (8%) | 29 (7%) | 0.50 | 0.83 | 0.50 |
| Other | 3 (1%) | 0 (0%) | 0 (0%) | 3 (1%) | NA | 0.60 | 0.56 |
| White | 450 (90%) | 43 (94%) | 34 (89%) | 373 (90%) | 0.51 | 0.97 | 0.41 |

**Bolded *p*-values** denote results significant at $p < 0.05$ .
**Note:** Between group comparisons are Mann-Whitney U and Wilcoxon rank sum tests.
**Abbreviations:** AD+MCI: Alzheimer's Disease or Mild Cognitive Impairment, APOE4+: Apolipoprotein E ε4 allele carrier, NDAN: Non-demented Alzheimer's neuropathology, SD: Standard Deviation.

### Linear Models Evaluating Group Differences in CBF and WMH

Results from models predicting CBF for NDAN and Controls relative to AD+MCI are shown in Table 3. Results are presented by region (frontal, temporal, parietal, occipital, limbic system, basal ganglia) and sorted based on regions associated with neurofibrillary tangle formation for Braak stages I-VI. Left hemisphere CBF in the middle cingulate gyrus (*p* < 0.05) and several of the temporal (all *p*s < 0.05), parietal (all *p*s < 0.05) and occiptal (all *p*s < 0.05) regions, as well as two of the limbic system structures (all *p*s < 0.05) distinguished NDAN from AD+MCI; CBF was significantly higher in NDAN, with largest effect sizes observed for the parietal (+22-30%) and the occipital (+29-34%) regions. Right hemisphere CBF in one frontal and several temporal, parietal and most occiptal (all *p*s < 0.05) regions also distinguished between Controls and AD+MCI. For the majority of the regions investigated, NDAN had greater CBF than Controls, but differences were not significant (all *p*s > 0.05).

**Table 3.**
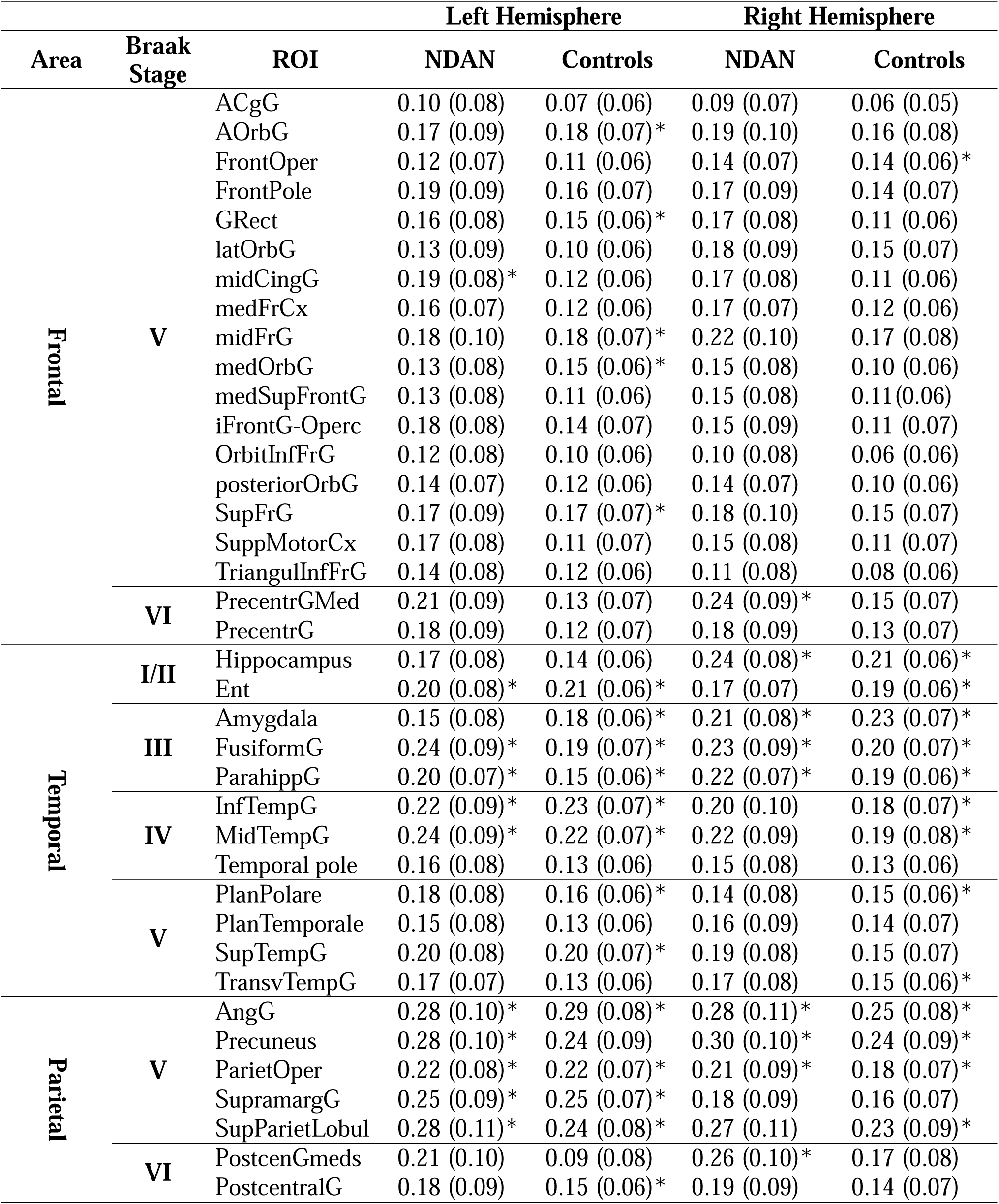
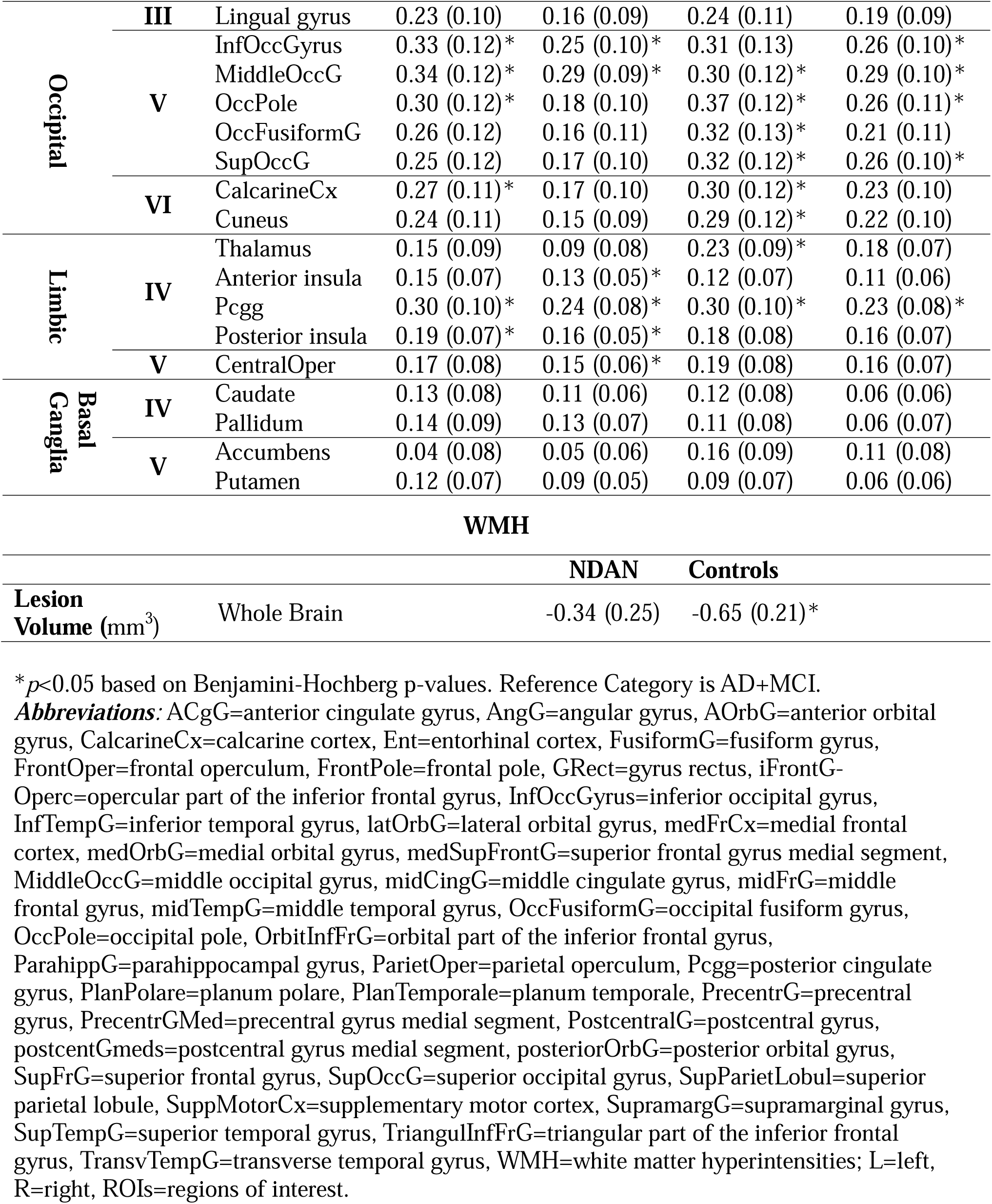
Results of regressions predicting cerebral blood flow (CBF) and white matter hyperintensities (WMH) by group. (adjusted % difference from AD+MCI (SE)).

We conducted sensitivity analyses adjusting for global cortical DVR to ensure that any oserved findings were not confounded by group differences in Aβ burden. We did not perform sensitivity analyses with tau-PET measures because the neocortical tau load (prevalence of Braak stage V or VI) did not differ significantly between groups. Our findings for the NDAN vs.

AD+MCI comparison remained largely unchanged, except for left middle cingulate gyrus, left entorhinal cortex, left parahippcampal gyrus, left inferior and middle temporal gyri, left occipital pole, and right amygdala, which, despite similar effect esimates, were no longer significant owing to small changes in standard error estimates and degrees of freedom. Analyses were also repeated after matching Controls to AD+MCI and NDAN 3:1 on 5-year age group and sex to ensure our findings were not a result of age or sex differences between groups; the findings persisted in the matched sensitivity analyses.

The results of linear regression model predicting WMH volumes for each group are displayed in the last row of Table 3. WMH volume was significantly lower in Controls (*p* < 0.002), but not NDAN (*p* < 0.05), compared to AD+MCI.

## DISCUSSION

NDAN had a significantly higher CBF compared to AD+MCI in the majority of gray matter regions where neurofibrillary tangles form throughout the course of AD, comparable to or slightly higher than the CBF in Controls. The most pronounced CBF differences between AD+MCI and NDAN were observed in posterior regions, specifically the occipital (+29-34% greater CBF in NDAN) and the parietal (+22-30%) cortices. Our findings are consistent with the existing literature which suggests that CBF, most notably in posterior regions, is associated with AD-related cognitive decline independent of core AD neuropathology.^40,47–49^ Furthermore, both functional abnormalities^57^ and brain atrophy ^58^ in posterior brain regions are associated with faster AD progression. Overall, the existing literature suggests that posterior regions, among the earlier areas of Aβ deposition^52^ but not typically affected by neurofibrillary tangles until late in AD,^33^ may have an important role in preserving cognition.

The observed CBF preservation in both NDAN and Controls compared to AD+MCI is reflected in the literature on vascular changes in AD. CBF decrease, among the earliest changes in AD progression and likely caused by contractile pericytes constricting capillaries, continues to be associated with cognitive decline over the course of the disease.^53^ Reduction in CBF is also closely associated with reduced glucose metabolism and synaptic failure,^7^ suggesting disruption in network connectivity, which is thought to reduce the efficiency of information processing and contribute to the cognitive deficits characteristic of AD.^48,54^ In addition, reduced posterior CBF is associated with cerebral amyloid angiopathy, which is common in AD and associated with faster cognitive decline.^48,55^ Given the documented relationships between CBF and cognition, preserved CBF in NDAN may help explain cognitive maintenance relative to AD+MCI when faced with AD neuropathology and may therefore be a promising prognostic marker of AD-related cognitive decline in A+T+ individuals.

Although not statistically significant, NDAN had a slight hyperperfusion relative to Controls in a majority of areas studied, potentially signaling a mounting response by NDAN against accumulating core AD neuropathology.^15,18^ Our findings are consistent with literature reporting region-specific hyperperfusion in AD typically associated with prodromal and asymptomatic stages of the disease that ultimately progresses to hypoperfusion as AD becomes symptomatic,^15–19^ but differs from other biomarker findings in NDAN, where typically NDAN exhibit biomarker levels intermediate to those of Controls and AD+MCI.^2^ Hyperperfusion in preclinical stages correlates with better cognition, particularly in individuals who do not carry genetic risk for AD.^56^ Nonetheless, our CBF findings suggest a potentially unique role for CBF in preserving cognition in NDAN and as such, warrant further study.

WMH burden was higher in AD+MCI than in Controls but did not differentiate NDAN from AD+MCI. However, a substantial literature links greater WMH burden to cognitive decline and AD progression^57^, including work by Brickman and colleagues showing that regional, particularly parietal, WMH are associated with AD.^58^ The apparent divergence between our CBF and WMH findings, and also the general literature, may reflect a proximal nature of regional CBF as a measure of cerebral perfusion and metabolic support, whereas global WMH burden is a cumulative and etiologically heterogeneous marker of white-matter injury. ^59,60^ WMH are commonly attributed to cerebral small-vessel disease, but evidence also supports contributions from AD-related neurodegeneration, cerebral amyloid angiopathy, and other nonvascular processes.^10^ Thus, region-speceific WMH load or lesion growth, may be more sensitive to vascular contributions to cognitive resilience than total whole-brain WMH volume. ^61^ Together, our findings suggest that WMH, although more prevalent in AD+MCI, do not explain the preservation of cognition in NDAN against the background of accumulating AD neuropathology at the whole-brain level.

The cross-sectional nature of this study necessitates future longitudinal investigation of CBF in individuals resilient to AD pathology. However, the significant CBF differences between NDAN and AD+MCI in many brain regions investigated suggest that cerebral hypoperfusion is a potential marker of future decline. Characterizing longitudinal changes in CBF between NDAN, AD+MCI, and Controls is therefore a priority for future work. For the purposes of comparison and interpretability, resilience was operationalized as a binary construct in the present analyses. We recognize, however, that resilience likely exists along a continuum reflecting both cognitive performance and the accumulation of neuropathology. Future studies with larger samples and sufficient statistical power should examine alternative thresholds and definitions of resilience, including approaches that account for specific cognitive domains and the regional distribution of tau pathology. Specifically, the NDAN and AD+MCI groups were smaller than the Control group, and replication in larger cohorts will be important to establish the robustness and generalizability of these findings. Finally, the cohort is enriched for Alzheimer’s disease risk and consists predominantly of educated, non-Hispanic White participants. These characteristics may limit the generalizability of our findings. Ongoing enrollment and data collection within the WRAP and WADRC cohorts will provide opportunities to address these limitations in future analyses. Nevertheless, CBF was consistently higher in NDAN in the majority of regions investigated and warrants further study as a marker of resilience.

Overall, our findings highlight the potential importance of preserved CBF as a biomarker of cognitive resilience in NDAN, who despite harboring AD pathology maintain cognitive function. The higher CBF in NDAN compared to AD+MCI, especially in posterior brain regions implicated in AD progression, suggests a protective role for vascular integrity in cognitive maintenance. Future longitudinal research with larger cohorts complemented by autopsy studies will be essential to confirm prognostic utility of CBF and clarify its role in cognitive preservation over the course of AD progression.

## AUTHOR CONTRIBUTIONS STATEMENTS AND DECLARATIONS

### Ethical Considerations

The University of Wisconsin Institutional Review Board approved all study procedures.

### Consent to Participate

Each subject provided signed informed consent before participation.

## Data Availability

All data produced in the present study are available upon committee approval.

https://wrap.wisc.edu/data-requests-2/

